# National statutory reporting: not even ticking the boxes? The quality of ‘Learning from Deaths’ reporting in Quality Accounts within the NHS in England 2017-2020

**DOI:** 10.1101/2022.07.12.22277525

**Authors:** Z Brummell, D Braun, Z Hussein, SR Moonesinghe, C Vindrola-Padros

## Abstract

**Introduction:** Regulation through statutory reporting is used in healthcare internationally to improve accountability, quality of care and patient safety. Since 2017 within the National Health Service (NHS) in England NHS Secondary Care Trusts (NSCTs) are legally required to report annually both quantitative and qualitative information related to patient deaths within their care within their publicly available Quality Accounts as part of a countrywide patient safety programme: The Learning from Deaths (LfDs) programme.

**Method:** All LfDs reports published between 2017 (programme inception) and 2020 were reviewed and evaluated through a critical realist lens, quantitatively reported using descriptive statistics and qualitatively using reflexive thematic analysis.

**Results:** In 2017/18 44% of NSCTs reported all six statutory elements of the LfDs reporting regulations, in 2019/20 35% of NSCTs were reporting this information. A small number of NSCTs did not report any parts of the LfDs regulatory requirements between 2017 and 2020. Multiple qualitative themes arose from this study suggesting problematic engagement with the LfDs programme, erroneous reporting accuracy and written communication.

**Conclusions:** The LfDs programme has to some extent reduced variation and improved consistency to the way that NSCTs identify, report and investigate deaths. However, three years into the LfDs programme the majority of NSCTs are not reporting as required by law. This makes the validity of National statutory reporting in Quality accounts within the NHS in England questionable as a regulatory process.

## Introduction

Statutory public reporting through the mandatory submission of both financial and non-financial information is used in healthcare internationally as a form of regulation, in an attempt to improve accountability and quality of care.[1,2] Within the National Health Service (NHS) in England, public reporting to improve quality has been a component of a National Quality Programme for nearly two decades.[3] There is mixed evidence whether publicly reporting quality and performance information improves outcomes for patients.[4]

The ‘Quality Account’(QA) is an annual report, publicly available on NHS Secondary Care Trusts (NSCTs) websites and submitted to the Secretary of State for Health and Social Care by end June each year; publication is mandated by law.[5,6] QAs provide data about the quality of NSCTs services. Some priorities for improvement are set and reviewed by individual NSCTs, others are statutory requirements.[7] All NSCTs, acute, specialist, mental health, community and ambulance services have the same requirements for inclusion in their QA. The Kings Fund (2011) reviewed QAs compliance with statutory requirements shortly after guidance introduction, they found significant variation between NSCTs quality measures, and the format of presentation. They recommended policy makers support organisations to ensure ‘clarity and consistency in the presentation of quality indicators’.[8]

Current legislation requires all NSCTs to annually report both quantitative and qualitative information related to patient deaths within their care, as part of a new programme – Learning from Deaths (LfDs).[9,10] LfDs arose in response to independent reviews of individual NSCTs which demonstrated a lack of systematic approach in the ways in which NSCTs become aware of, investigate and share learning from deaths and a lack of meaningful change occurring in response to unexpected deaths.[11,12] The NHS National Quality Board (a forum of senior clinicians from key national NHS oversight organisations, including the Care Quality Commission (CQC)) published ‘national guidance on Learning from Deaths’, describing mortality governance, engagement with bereaved families and the requirement of LfDs reporting in QAs.[10] The LfDs programme was not piloted, the logic model was not clear and evaluation was not built into the programme.

Lalani and Hogan (2021) in their narrative account of the key drivers in the development of the LfDs programme noted that some NSCTs have taken on the LfDs programme as a ‘tick-box’ exercise.[13] This study therefore analyses LfDs reports to understand whether NSCT reporting in their QAs matches statutory requirements and to review the quality of reporting. A qualitative analysis seeks to understand any difficulties around reporting. This study does not analyse the ‘Learning’ or ‘Actions’ or ‘assessment of the impact of the actions’ arising through the NSCT reviews, this will be discussed in a separate paper.

## Method

This is a qualitative and quantitative study evaluating the quality of statutory reporting for an NHS improvement programme. The analysis is undertaken using a critical realist lens.[14]

We undertook a secondary source document analysis using quantitative counts and descriptive statistics. For qualitative analysis, we used thematic analysis of all LfDs reports in QAs from NSCTs between 2017 – 2020. Ambulance trusts were excluded as they were not required to report in 2017/18.[15,16]

Reflexive Thematic Analysis (TA) as described by Braun and Clarke (2021) has been used for its flexible, yet structured approach to find “shared meaning” in the data while enabling researcher consideration of their own impact on data interpretation.[17] Reflexive TA in this study enables evolution of and contextualisation of the causal mechanisms affecting the ability of NSCTs to report LfDs. The authors have purposefully used quantitative and qualitative approaches, despite the tensions arising, to provide a fuller explanation of NSCTs empirical reporting, to engage with frontline staff and policymakers (who embrace quantitative research) and to reveal underlying deeper causal mechanisms negatively impacting LfDs reporting.

We undertook analysis of LfDs as set out in the 2017 amendment to the NHS 2010 quality account regulations: this involved review of compliance of reports against regulation numbers 27.1 to 27.6 (table 1).[9] Where NSCTs did not fully report we sought to understand why this may have been the case from comments within the QA itself. In addition, we reviewed LfDs reports for basic errors in quality (such as grammatical and typographical mistakes). Only data found within the NSCTs QA was included in the analysis. Quantitative analysis was undertaken and reported using descriptive statistics. The qualitative methodology used was a process of data familiarisation (reading each report twice on separate occasions), systematic data coding, generating initial domains deductively, then developing multi-dimensional themes inductively through active engagement and immersion in the data, looking both at what was present and what was absent. Finally, these themes were refined by sense checking with the original reports and discussion with the other authors.

**Table 1:** NHS Quality Accounts LfDs Regulations [9].

| **Regulation number** | **Summary of regulatory requirement** |
| --- | --- |
| **27.1** | The number of patients who have died during the annual reporting period |
| **27.2** | The number of the deaths (in 27.1) that have undergone a case record review or investigation |
| **27.3** | An estimate of the number of deaths in 27.2 which the provider judges to be more likely than not to have been due to problems in care, with explanation of method to assess this |
| **27.4** | What the provider has learnt from reviews/investigations in relation to deaths (in 27.3) |
| **27.5** | A description of the actions the provider has taken or will take in response to what they have learnt |
| **27.6** | An assessment of the impact of the actions (from 27.5) |

Every LfDs report was reviewed by the primary reviewer (ZB) twice, on separate occasions to ensure full data capture. 10% of reports from 2018/19 and 2019/20 were identified by random number generation and reviewed independently by a second reviewer (ZH) to ensure accuracy and reliability. In the case of disagreement, ZB re-reviewed the LfDs report for clarification. Reflexive TA was predominantly undertaken by ZB, with input from the other authors at the final stage.

Data were captured in Microsoft excel (V.16.15). This study has been reported using Standards for Reporting Qualitative Research.[18]

### Reflexivity

Reflexivity was undertaken alongside the research methodology, as described by Trainor & Bundon, 2021.[19] For additional information see supplemental page 1.

### Patient and public involvement

This study forms part of a larger programme of work overseen by a public and relatives steering group to improve relevance from the perspective of those affected by deaths in healthcare and to reduce healthcare researcher bias. The steering group have been involved in the planning, design, development of conclusions and paper authorship. The reporting of PPI has been undertaken using guidance for reporting involvement of patients and the public 2— short form.[20]

## Results

### Quantitative Analysis

The number of NSCTs is reducing year on year due to mergers of NSCTs: 222 NSCTs in 2017/18, 217 NSCTs in 2018/19, 213 NSCTs in 2019/20.

NSCTs who reported all six statutory elements (table 1) of the LfDs reporting framework:

- 2017/18: 98 out of 222 (44%)
- 2018/19: 109 out of 217 (50%)
- 2019/20: 75 out of 213 (35%)

NSCTs who did not report any parts of the LfDs regulatory requirements:

- 2017/18: Two NSCTs
- 2018/19: Three NSCTs
- 2019/20: Eleven NSCTs (9 QA published, but no LfDs report, 2 no QA published)

The total number of deaths reported by individual NSCTs:

- 2017/18: 3 deaths to 7756 deaths (mean 1341, median 1211, range 7753)
- 2018/19: 2 deaths to 7399 deaths (mean 1301, median 1154, range 7397)
- 2019/20: 1 death to 7939 deaths (mean 1438, median 1219, range 7938)

The number of case record reviews or investigations undertaken relative to the number of patient deaths in individual NSCTs varied:

- 2017/18: between 0.2% and 100% of deaths (mean 43.7, median 36.5, range 99.8)
- 2018/19: between 0.9% and 100% of deaths (mean 43.7, median 37.1, range 99.1)
- 2019/20: between 0.7% and 100% of deaths (mean 45.1, median 36.7, range 99.3)

The number of deaths which the provider judged to be more likely than not to have been due to problems in care

- 2017/18: between 0 and 13% (mean 0.7, median 0.2, range 13)
- 2018/19: between 0 and 8% (mean 0.005, median 0.002, range 8)
- 2019/20: between 0 and 100% (mean 0.5, median 0.2, range 100)

In 2019/20 one NSCT, reporting 1 death in total, with that death judged more likely than not due to problems in care, therefore skewing the data.

Lack of reporting any deaths judged to be more likely than not to have been due to problems in care (excluding NSCTs who didn’t report at all)

- 2017/18, 20 NSCTs did not report
- 2018/19, 18 NSCTs did not report
- 2019/20, 19 NSCTs did not report

Reasons given for not reporting included:

- ‘*data collection challenges’*
- *‘unable to provide a reliable figure’*
- *‘we do not carry out investigations with a view to determining whether the death was wholly or partly due to problems in the care provided’*
- *‘currently no research base on this for mental health services and no consistent accepted basis for calculating this data’*

Reasons for not reporting are discussed further within the qualitative reflexive thematic analysis.

NSCTs who published a LfDs report, but did not report any lessons learnt from deaths

- 2017/18: 25 out of 220 NSCTs (11%)
- 2018/19: 23 out of 214 NSCTs (11%)
- 2019/20: 29 out of 202 NSCTs (14%)

NSCTs who published a LfDs report, but did not report any actions taken as a result of learning

- 2017/18: 30 out of 220 trusts (14%)
- 2018/19: 24 out of 214 trusts (11%)
- 2019/20: 25 out of 202 trusts (12%)

NSCTs who published a LfDs report and provided information about an assessment of impact of actions:

- In 2017/18: 105 out of 220 trusts (47%)
- In 2018/19: 123 out of 214 trusts (57%)
- In 2019/20: 79 out of 202 trusts (39%)

### Qualitative Reflexive Thematic Analysis

#### ‘This programme isn’t for us’

It was clear from the reports particularly in 2017/18, that some NSCTs did not believe the guidance was written for or applied to them. The similarity with some of the wording in the LfDs reports could indicate collaboration between NSCTs about how to approach elements of the regulation deemed more controversial, particularly reporting the number of death more likely than not due to problems in care. This was especially the case for Mental Health and Community NSCTs: *‘there was no nationally agreed definition for mental health services or community health services with regard to what constitutes a death from problems in care and therefore this data was not reported’*.

A small number of NSCTs reported defensively, for example: ‘*Data presented within this section is for learning within the Trust and is not comparable with any other trusts (Acute, Community and Mental Health) published data, it should not be used to provide organisational benchmarking or presented as comparators in any onwards reports’*.

Another less common occurrence was wording demonstrating annoyance: ‘*NHS England did not make any central training resource available to roll out training on how the mortality review tool should be implemented by mental health trusts across the country and to spread learning from it both locally and nationally’*.

One NSCT did report in 2017/18 and 2018/19, but subsequently have not provided a LfDs report in 2019/20 (or in 2020/21) despite commenting in their QA that they have a *‘Detailed learning from deaths programme in place’*.

Some NSCTs did report a death or deaths due to problems in care, but do not report any learning or actions as a result of this; none explain why this is the case. For example: One NSCT reported 11 deaths more likely than not due to problems in care, however in the lessons learnt and actions sections they have stated ‘*N/A’*.

The statutory requirement to report the number of deaths due to problems in care appears to be one of the most difficult for NSCTs to report on. As can be seen from the quantitative data, many NSCTs reported zero deaths or did not report the number of deaths due to problems in care. One NSCT describes being ‘*unable to provide a reliable figure for the number of deaths in the reporting period which were judged more likely than not to have been due to problems in the care’* because the methodology they are using does *‘not allow the calculation of whether a death has a greater than 50% probability of being avoidable*’ and add to this that this methodology provided by the Royal College of Physicians does not *‘endorse the comparison of data’* between NSCTs.

#### ‘A partial truth’

A very small number of NSCTs make approximations in their LfDs reports related to deaths, one NSCT described that the ‘*mortality review process identified* ≤*5 deaths that were judged to be more likely than not to have been due to problems in the care provided to the patient*.’ Another NSCT does not give an exact number of total deaths, but instead states that ‘*around 3000 inpatients die in our hospitals every year’*.

One NSCT describes no deaths due to problems in care, but also note receipt of a Prevention of Future Death (PFD) report from the coroner within the same QA. The coroner issued a PFD report in describing problems in care where actions could be taken.[21]

Another NSCT reporting zero deaths due to problems in care in their 2018/19 LfDs report also report within the same QA that there were 23 patients who came to severe harm or death due to patient safety incidents in 2018/19. While it is possible that all these incidents caused severe harm (and not death) this is not stated.

#### ‘Not speaking the same language’

Reporting is heterogenous. NSCTs have reviewed and investigated with varying thoroughness and with differing methodologies. Some NSCTs have not reported data for the same time periods (for example reporting annual data from Jan to Dec instead of April to April). Some NSCTs reported incomplete data for total number of deaths and/or reviews/investigations (for example only reporting for 9 months of the year). NSCTs have assessed deaths in scope for inclusion variably, with variable inclusion and exclusion of Emergency Department patients, Children, Stillbirths and outpatients.

Reviewers and investigators are also not standardised and may not always be suitably qualified to undertake a review: ‘*Investigation of patient’s complaint of pain in his neck should have been investigated earlier, although it is beyond the experience of the reviewer as to how common this complaint is after tracheostomy’*.

Other elements of reporting are also difficult to compare, for example what one NSCT thinks is assessment of impact is not what another considers to be the case. Some NSCTs have focused heavily on the process around LfDs for example which patients are in scope for review, but provide very little detail about the incident.

#### ‘Errors in written communication’

Errors in written communication are a frequent reoccurrence across a large number of NSCTs LfDs reports. Many of these mistakes in isolation could be considered minor, but together account for a significant number of mistakes in a public facing document about people who have died.

Some of these errors could be deemed careless typographical and/or mathematical errors, for example: misplacing a decimal point, thus completely change the scale of the data. Another relatively common error is not providing a heading or title for the LfDs report, or for tables or graphs. Or mistakenly writing the wrong financial year within the report. Or duplication of information. Another occasional problem is contradictory information in the same report.

Occasionally sentences within the LfDs reports are difficult to interpret or don’t make sense, due to grammatical errors: in one NSCT LfDs report they state: ‘*In 2018-19 specialty Mortality & Morbidity meetings the quality of case discussions has been improved through the additional collective judgement of the overall quality of care using the NCEPOD grading tool’*

## Discussion

This study demonstrates a significant lack of LfDs reporting in QAs between 2017 and 2020 and a signal that many NSCTs did not fully understand the statutory requirements or did not think they applied to them (‘This programme isn’t for us’). The overall reporting for all six statutory elements of the LfDs reporting framework (table 1) had reduced from 44% in 2017/18 to 35% by 2019/20. This reduction is in contrast to expectations three years after the introduction of the programme.[22] It is possible that the COVID-19 pandemic, affecting NSCTs by March 2020, could account for this, but this seems unlikely to be the whole story since the QA reporting year ended in April 2020. When we previously reported on the quality of 2017/18 LfDs reporting, we noted that reporting variation may be due to differences in interpretation of the guidance and statutory requirements; however three years should be sufficient time to get to grips with reporting requirements.[23] The incentive to report appears clear: to demonstrate quality of care and safety for patients; however, the penalty for not reporting is less clear, there is no direct financial penalty for an NSCT not reporting LfDs. The only sanction facing NSCTs is that they may be notified to take action following CQC inspections.[24] And the evidence that external inspections actually improve NSCT performance is unclear.[25]

There remains wide variation in the total number of deaths reported by NSCTs. This may in part be accounted for by the different types of NSCT, where specialist NSCTs will have fewer deaths and community NSCTs could have thousands of deaths if outpatients are included. However, the wide variation in the number of case record reviews or investigations undertaken relative to the number of patient deaths in individual NSCTs, is less easily explained and was a concern raised by the CQC in 2016.[12] There has been improvement in the standardisation of NSCT case note review methodology, with the majority of NSCTs (particularly acute NSCTs) using Structured Judgement Review (SJR) methodology by 2020.[26] Mental Health NSCTs were however less likely to be using the SJR methodology and overall appeared to be less engaged with the LfDs programme.

Some NSCTs (between 11 and 14% depending on the year of analysis) did not report any lessons learnt from deaths; some reported zero deaths more likely than not due to problems in care and therefore were not strictly required to report lessons learnt. There were also a small number of NSCTs who reported deaths more likely than not due to problems in care but did not go on to report lessons learnt. As one example, an acute NSCT reported 11 deaths due to problems in care (2017/18), but then reported ‘N/A’ in learning and actions. This NSCT in their last CQC inspection (2019) was reported as being ‘Good’ overall and ‘as ‘Requires improvement’ for safety, LfDs was not mentioned in the CQC report and it is not clear if the NSCT understood LfDs statutory reporting requirements.[27] As discussed in ‘This programme isn’t for us’ several NSCTs raised concerns about the requirement to report the number of deaths due to problems in care. This concern was also raised by NSCT representatives at NHS England LfDs guidance development days in 2017, where problems with the term ‘avoidability’ were raised (event attended by ZB and DB), due to the negative impact on NSCT reputation.

Written communication such as that in the QA is used to both give information and to persuade the reader that the NSCT is safe and that quality healthcare is being provided. This communication should be clear, complete, accurate, concise and honest. A lack of attention to detail with regards to written communication, could legitimately raise concerns about data accuracy and could potentially make the general public concerned about a lack of attention to detail in clinical care. With this in mind ‘errors in written communication’ have greater significance than might be initially apparent. The small number of NSCTs highlighted in ‘a partial truth’ with approximations for the number of deaths and clear contradictions in reporting within the QA provide similar but potentially even greater concerns.

It is not usually apparent who has collected, collated and compiled the data within a QA. However, all QAs are ‘*signed by the responsible person for the provider that to the best of that person’s knowledge the information in the document is accurate’*. This is usually someone from the NSCT Board, often the Chief Executive and/or the Chair. NSCTs are also required to share their QAs with Commissioners, to their local Healthwatch and to Overview and Scrutiny Committees.[28]

### Lack of consensus on how to measure quality

The measurement of quality through outcome data and the intention to use this to improve health services is commonplace in healthcare internationally. There is little consensus on which measures and how many of these indicators are best used to assess quality.[29] Data collection, collation and presentation for QAs is resource intensive, but if deemed a statutory requirement should be provided in its entirety by NSCTs, even if viewed a ‘tick-box’ exercise.[30] QA data can be utilised to improve quality of care, accountability and patient choice.[31] However when that data is not provided and there is no recourse for this, it makes a mockery of the whole system. It is possible that the burden of regulation has become too much for some NSCTs, particularly where there is little feedback from regulators about how the data is used or if it is actually useful.[32]

Published research reviewing or analysing the content of NSCT QAs is very limited. Jones et al (2017) found that NSCTs with high Quality Improvement (QI) maturity had QAs with clear internally driven priorities, in contrast to those with low QI maturity.[33]

### The use of mortality data

The use of data related to hospital mortality to assess performance (and safety) has been a contested issue for many years. The reasons behind this include concerns about heterogeneity of data (even with risk adjustment) between NSCTs and the accuracy of data. Lilford and Provonost (2010) strongly argued that the use of mortality rates to assess performance for regulatory purposes was wrong, due to the lack of specificity provided by this data and unfair penalisation. They raised concerns that regulation associated with mortality rates could lead to falsification of reporting or even ‘overly aggressive care’ and that identification (and subsequent investigation) of poorly performing NSCTs could detract from improvements across all NSCTs.[34] NSCTs should however be aware of how many people died in their care and have robust processes in place for families (and staff) to highlight concerns that a death might have been due to problems in care. All people will die and most deaths cannot be prevented. But, where a death has occurred due to problems in care, an open and honest investigation is required, actual learning about what the problem was and why it happened must occur, improvement action is needed and processes must exist to evaluate the impact of any interventions. There is a healthcare (and perhaps societal) cultural reluctance to accept death. This reluctance may have a part to play in why some NSCTs have difficulties in engaging with LfDs.[35]

### The impact of public reporting

The evidence that NSCT QI, public reporting or even legislation improves health outcomes or patient safety is sparse.[4, 36, 37] The costs associated with healthcare policy and interventions are sometimes overlooked in lieu of the possibility of a quick fix.[38] This in addition to a lack of consideration for the complexity and importance of support for implementation may prevent health policy success.[39] Evaluation of countrywide QI programmes such as the LfDs programme are not commonly undertaken. Evaluation should be built into these programmes at the planning stage and should form an essential component of all health policy.[40] There is limited understanding about the effectiveness of QI interventions in supporting failing organisations. Underperforming NSCTs have less time and resources available for new QI programmes, therefore consideration for implementation and what support might be needed should be given.[41, 42]

### Next steps

For the LfDs programme, we would recommend robust regulatory reporting oversight in addition to CQC inspections.

## Conclusions

When a patient dies in an NSCT due to problems in care this death must be identified, investigated and where possible actions taken to prevent future deaths. Accountability and an understanding of human fallibility must be balanced. The LfDs programme has to some extent reduced variation and improved consistency to the way that NSCTs identify, report and investigate deaths. However, three years into the LfDs programme the majority of NSCTs are not reporting as legally required and current evidence that LfDs reporting has improved patient safety remains elusive.

Public statutory reporting even if done well may not improve quality of care or patient safety; therefore, further research is needed to assess the value of QAs. With the LfDs programme, the inability of many NSCTs to ‘tick the box’ of basic statutory requirements limits successful evaluation. The findings from this study bring into question the validity of statutory reporting in QAs as a regulatory process.

## Data Availability

All data used is publicly available from NHS Secondary Care Trust websites in England

## Acknowledgements

We would like to acknowledge the work of the Learning from Deaths: Learning and Action (LfDLaA) Public and Relatives steering group in this research. Patient and public involvement in this research was supported by the NIHR UCL Biomedical Research Centre.

## Supplemental page 1

### Reflexivity detail

The primary reviewer (ZB) is a frontline clinician, who felt embarrassment and disappointment during the collection and analysis phases of this research, by the lack of attention to detail, disregard for undertaking statutory requirements and often indifference demonstrated by NSCTs. It appeared to ZB that some NSCTs and those ‘signing off’ LfD reports, see the LfDs programme as another regulatory requirement and not as an opportunity to improve care for future patients. ZB understands the challenges of working in frontline healthcare and appreciates that NSCTs have many competing programmes and issues to manage. ZB shared these reflections and concerns during meetings with the other authors, this process of team-based discussion helped identify assumptions and cross check interpretations.

## Supplemental page 2

### Quality Accounts with direct quotes in paper

- Lancashire and South Cumbria Quality Account 2019/20
- Lincolnshire Partnership Quality Account 2018/19
- Black Country Quality Account 2019/20
- London North West Quality Account 2019/20
- Poole Hospital Quality Account 2017/18
- Royal Devon and Exeter Quality Account 2018/19
- City Hospitals Sunderland Quality Account 2018/19
- Barts Health Quality Account 2017/18
- Coventry and Warwickshire Partnership Quality Account 2018/19
- South Tees Quality Account 2019/20
- PAH Quality Account 2019/20
- Sherwood Forest Quality Account 2018/19

